# Cellular and humoral responses to SARS-CoV-2 vaccination in immunosuppressed patients

**DOI:** 10.1101/2021.12.03.21267250

**Authors:** Dinesh Mohanraj, Samuel Baldwin, Satbeer Singh, Alun Gordon, Alison Whitelegg

## Abstract

**Objective:** SARS-CoV-2 vaccinations have demonstrated vaccine immunogenicity in healthy volunteers, however, efficacy in immunosuppressed patients is less well characterised. Subsequently, there is an urgent need to address the impact of immunosuppression on vaccine immunogenicity.

**Methods:** Serological, T-cell ELISpot, cytokines and immunophenotyping investigations were used to assess vaccine responses (either BNT162b2 mRNA or ChAdOx1 nCoV-19) in double-vaccinated patients receiving immunosuppression for renal transplants or haematological malignancies (n=13). Immunological responses in immunosuppressed patients (VACC-IS) were compared to immunocompetent vaccinated (VACC-IC, n=12), unvaccinated (UNVACC, n=11) and infection-naïve unvaccinated (HC, n=3) cohorts. All participants, except HC, had prior COVID-19 infection.

**Results:** T-cell responses were identical between VACC-IS and VACC-IC (92%) to spike-peptide (S) stimulation. UNVACC had the highest T-cell non-responders (n=3), whereas VACC-IC and VACC-IS both had one T-cell non-responder. No significant differences in humoral responses were observed between VACC-IC and VACC-IS, with 92% (12/13) of VACC-IS patients demonstrating seropositivity. One VACC-IS failed to seroconvert, however had detectable T-cell responses. All VACC-IC participants were seropositive for anti-spike antibodies. Furthermore, both VACC-IS and VACC-IC participants elicited strong Th1 cytokine response with immunodominance towards S-peptide. Differences in T-cell immunophenotyping were seen between VACC-IS and VACC-IC, with lower CD8^+^ activation and T-effector memory phenotype observed in VACC-IS.

**Conclusion:** SARS-CoV-2 vaccines are immunogenic in patients receiving immunosuppressive therapy, with responses comparable to vaccinated immunocompetent participants. Lower humoral responses were seen in patients treated with B-cell depleting therapeutics, but with preserved T-cell responses. We suggest further work to correlate both protective immunity and longevity of these responses in both healthy and immunosuppressed patients.

## INTRODUCTION

In late 2019, identification of a novel coronavirus, severe acute respiratory syndrome coronavirus 2 (SARS-CoV-2), was described as the causative pathogen of a pneumonia outbreak, known as coronavirus-induced-disease-19 (COVID-19) [1]. What emerged as a local outbreak in Wuhan, China, rapidly progressed into a global pandemic of acute respiratory syndrome evoking mass morbidity, mortality and significant socio-economic turmoil [2]. Currently, mass vaccination programmes, utilising regulatory-approved vaccines, remains the best way to prevent viral transmission [3,4], severe disease, death [5,6] and overwhelming the already stretched healthcare services.

Currently, four vaccines have been approved by the European Medicines agency [7,8], demonstrating satisfactory safety and immunogenicity. However, these pre-authorisation trials were performed on healthy individuals and excluded immunosuppressed patients as they are poor responders to vaccines [9, 10]. Consequently, ambiguity regarding vaccine efficacy in patients on immunosuppression prevails. Moreover, immunosuppressed patients, such as kidney transplant recipients, have been considered as clinically vulnerable to SARS-CoV-2 infection, which is supported by both population-based and registry-based studies which illustrate these patients experience significant rates of hospitalisations, severe disease and death [11–13]. In view of this, characterising vaccine-induced immune responses is crucial for understanding their protective immunity and formulating optimal immunisation regimes.

Both natural infection and SARS-CoV-2 vaccination induce spike protein specific antibodies with neutralising activity [8,14]. Nevertheless, the longevity and duration of such humoral protection is unclear, with several studies demonstrating waning antibody-levels over time [15]. In contrast, several findings have highlighted the role of long-term SARS-CoV-2 T-cell responses [16]. Effective cellular immune responses were attributed to mild-COVID-19 [17], alongside development of robust SARS-CoV-2 specific T-cells which were detected 6-8 months post-infection [18]. Moreover, both mRNA and adenoviral vaccines stimulated potent T-cell mediated responses in study-participants [19,20]. Furthermore, long-term duration of protective T-cell responses were identified against SARS-CoV, whereas no antigen-specific B-memory cells or antibodies were detected 6 years post-infection [21]. As such, when assessing vaccine immunogenicity, it is critical to assess both humoral and cellular responses. Such evaluation is of greater importance in immunosuppressed cohorts, as there is an urgent need to understand the impact of immunosuppression on the efficacy of SARS-CoV-2 vaccinations.

To address this knowledge gap, we assessed the effect of immune deficiency in vaccine specific responses in a cohort of immunocompromised patients. SARS-CoV-2 vaccine responses were assessed in adult-vaccinated kidney transplant patients, or those with haematological malignancies. Here we provide a detailed description of the cellular and humoral responses, following two doses of either mRNA or adenoviral-vector SARS-CoV-2 vaccines. Unlike current studies examining findings of early post-vaccine period, we define details of their most current response (median time: 115 days post-second dose). Based on our findings, we were able to conclude that these immunosuppressed patients produced an immunological response, to SARS-CoV-2 vaccines, which were comparable to healthy vaccinated participants. Our findings warrant further investigation to determine correlation between such observed responses with protective immunity and longevity, within this clinically vulnerable cohort. Moreover, such studies are imperative for the understanding of cellular responses towards the continual emergence of SARS-CoV-2 variants of concerns.

## METHODS

### Study design

This study was approved by the institutional research review board of Portsmouth Hospital University NHS Trust and ethical approval was obtained from a national ethics committee (London-City and East research ethics committee, IRAS: 291009). This study was registered under National Institute of Health Research (NIHR) portfolio (CPMS ID: 48275). The study was conducted in accordance with principles of Good Clinical Practice. All enrolled participants were aged ≥18 or over. Participants were assessed for study eligibility by providing a clinical history. Before enrolment, all participants provided written informed consent.

All recruited participants were convalescent donors, except for healthy controls who were COVID-19 infection-naïve and unvaccinated. All convalescent individuals had prior positive real-time polymerase chain reaction (RT-PCR) results before study enrolment. Participants were stratified into the following cohorts: healthy unvaccinated COVID-19-infection naïve (HC, n=3), unvaccinated (UVACC, n=11) and vaccinated immunocompetent healthcare workers (VACC-IC, n=12) and vaccinated immunosuppressed participants (VACC-IS, n=13). VACC-IS participants were put forward to the study by their respective clinicians, whereas the remaining participants were enrolled through hospital communications.

Blood samples were collected upon enrolment, which were taken in heparinized, EDTA and SST-collection tubes. Samples were processed within 8h of venepuncture. SST-collection tubes were centrifuged at 2000g for 10 minutes for collection of serum. Collected serum was stored at −20°C until SARS-CoV-2 serological assays were performed. EDTA tubes were used to perform a full-blood count using the DxH Haematology Analysers (Beckman Coulter). Heparinized tubes were processed for peripheral blood mononuclear cells (PBMCs) collection as described below for ELISpot analysis.

### T-cell ELISpot

SARS-CoV-2-specific T-cell responses were identified using the T-Spot *Discovery* SARS-CoV-2 (Oxford Immunotec) according to manufacturer’s instructions. In brief, leucosep tubes (Oxford Immunotec) were used to isolate PBMCs from lithium-heparinised whole blood. A total of 2.6×10^6^ PBMCs were plated into each individual well of T-spot plate. Each well is coated with one of the four different SARS-CoV-2 structural peptides; Spike (S1) protein, nucleocapsid (NC), membrane (MN) protein, and homology (segments of similar sequences which were eliminated from NC and MN panel). Negative and positive controls (phytohaemagglutinin) were used to control for cellular contamination and functionality, respectively. PBMCs were incubated overnight (37°C, 5% CO2) for 20 hours and IFNϒ-secreting SARS-CoV-2 specific T-cells were detected by using an automated plate reader (Autoimmun Diagnostika-ASK JM). IFN? secreting SARS-CoV-2 T-cells were reported as spot forming units (SFU) per well.

### SARS-CoV-2 cellular immunophenotyping

Following SARS-CoV-2 peptide stimulation in ELISpot plate, harvested PBMCs were counted to 2.0×10^6^. Counted PBMCSs were resuspended in FACS buffer (phosphate buffered-saline with sodium chloride, Beckman Coulter) and stained with a tetra 1 backbone (CD45-FITC, CD3-PC5, CD4-RD1, CD8-ECD (Beckman Coulter)), added with another two fluorochrome-labelled monoclonal antibodies (mAbs), HLA-DR-PC7 and CD38-Alexa-fluor 750, as markers for T-cell activation. PBMCs were incubated at room temperature in dark for 15 minutes and then resuspended with 250µl FACS buffer. Up to 2.0×10^6^ PBMCs were counted using a 10-laser Navios Flow cytometer (Beckman Coulter).

For T-cell subset analysis, harvested PBMCs following S-peptide stimulation were counted as outlined above. PBMCs were then stained with Duraclone IM T-cell panel (Beckman Coulter, Miami, FL). Signals from the following different fluorochrome-labelled mAbs were obtained; CD45-Krome Orange, CD3 APC-A750, CD4-APC, CD8-AF700, CD27-PC7, CD57-Pacific Blue, CD279 (PD1)-PC5.5, CD28-ECD, CD197 (CCR7)-PE and HLA-DR-FITC. PBMCs were incubated at room temperature in dark for 15 minutes and then resuspended with 500µl FACS buffer. Up to 2.0×10^6^ PBMCs were counted using a 10-laser Navios Flow cytometer (Beckman Coulter).

### Representation of high-dimensional flow cytometry

Flow cytometric t-distributed stochastic neighbor embedding (tSNE) and FlowSOM analysis were performed using Cytobank (http://premium.cytobank.org). For surface T-cell activation marker expression, analysis was performed using the above outlined markers. CD3^+^ gated events from individuals within each cohort were collected and concatenated into a single file. Data from 103,311 CD3+ gated events, from all cohorts per peptide, were exported as flow cytometry standard (FCS) files using Kaluza 2.1 software (Beckman Coulter, Miami, FL). Then, data from 4453 CD3+ events, with the following settings: 1,000 iterations, perplexity 30, and theta 0.5, subsampling equal each cohort was used to generate tiSNE analysis. FlowSOM was performed using above outlined markers, which were performed individually with gated CD4^+^ and CD8^+^ populations from tiSNE analysis per cohort. The following parameters were used to conduct FlowSOM analysis: number of clusters: 225; number of metaclusters: 15; iterations: 10; and hierarchical consensus clustering method was used.

For T-cell subset, analysis was conducted using above outlined markers. S-peptide stimulated CD4^+^ and CD8^+^ gated events from each individual were concatenated into a single file per cohort; VACC-IC CD4: 83,612 events; VACC-IC CD8: 35,721; VACC-IS CD4: 60,932; VACC-IS CD8: 16,397 events. All CD4 and CD8 events, per cohort, were used to conduct tiSNE analysis with aforementioned parameters. For both T-cell activation and T-subset analysis, heat maps were used to report statistical phenotypic changes in marker expression within CD4 and CD8 populations per cohort.

### Serological testing

Serum was tested for antibodies to Spike (S) protein using the Binding site Anti-spike IgG/A/M ELISA assay according to manufacturer’s instructions. Result outcomes are reported as positive or negative with a threshold index-value of ≥1.0. Samples with optical density greater than top-standard of curve were reported as >4.00 index value.

### Th1 cytokine profiling

Th1 cytokine responses (IL-6, TNF, IL-1β, IL-10) were measured in supernatant derived from PBMC stimulation with SARS-CoV-2 peptides within ELISpot plate. 20µl of supernatant was collected and stored in −80°C until analysis was conducted. IFN-γ was not tested as supernatant was derived from ELISpot plate which captures IFN-γ secretion. Cytokine responses were measured using Multiplex assays as performed by the Clinical Immunology laboratory at Addenbrooke’s Hospital, Cambridge.

### Statistical analysis

Statistical analysis was conducted using Prism V9.0 (GraphPad Software, San Diego, California, USA). Unless otherwise stated, all data are reported as median with IQR. Where appropriate, Kruskal-Wallis test with Dunn’s post-hoc comparison test was performed to assess differences between >2 groups. Two-sided Mann-Whitney was used to assess perform differences between 2 groups. *P*<0.05 unless otherwise stated. Other details, if any, for each investigation are provided within relevant figure legends. \**P*<0.05, \*\**P*<0.01, \*\*\**P*<0.001, \*\*\*\**P*<0.0001

## RESULTS

### Study participant characteristics

A total of 39 participants were recruited into the study and stratified into appropriate cohorts based on their clinical characteristics. In unvaccinated cohort (UNVACC, supplemental table S1), the median age of participants was 37 years (IQR: 31-47), male-to-female ratio was 3:8, with 81.8% of participants from white-British ethnicity. Co-morbidities of hypothyroidism (n=1), stroke (n=1) and sleep apnoea (n=2) were reported in this cohort. The reported time between positive RT-PCR result and study enrolment was 160 days (IQR: 145-165). Nine UNVACC participants (81.8%) were classified as ambulatory mild disease [22], based on reported signs and symptoms during active SARS-CoV-2 infection. Two participants were hospitalised requiring oxygen therapy (non-invasive ventilation) and treated with dexamethasone, which as shown by the RECOVERY trial (NCT: NCT04381936), lowered mortality in hospitalised adult COVID-19 patients.

Twelve participants were stratified as vaccinated immunocompetent (VACC-IC, supplementary table 2) with a median age of 45 years (IQR: 30-53) and male-to-female ratio of 1:5. Five VACC-IC participants reported co-morbidities of depression (n=2) and mild asthma (n=3); none of these participants, alongside remainder of VACC-IC cohort, were treated with immunosuppressive therapies. Time reported between positive RT-PCR result and study enrolment was 175 days (IQR: 143-431), with all twelve participants classified as having mild COVID-19 disease, and double-vaccinated with BNT162b2 vaccine. Median time between receiving the second vaccine dose and study enrolment was 112.5 days (IQR: 87.2-153).

Thirteen immunosuppressed patients (VACC-IS), with a median age of 61 years (IQR: 55-65), were recruited. Male-to-female ratio was 7:6, with all patients from a British-white ethnic background. Clinical characteristics and immunosuppressive regimes are summarised in Table 1. Seven patients (53.8%) had end-stage renal disease and received renal transplantation with an average of 2043 days (6.5 years) prior to study enrolment. Four patients were diagnosed with haematological malignancies, whilst two patients had autoimmune disorders. Twelve patients had further co-morbidities, of which all were high-risk for severe COVID-19. All VACC-IS were on immunosuppressive treatments, with 61.5% and 46.1% managed on Mycophenolate mofetil and Tacrolimus, respectively, and five patients receiving B-cell depletion therapy (Rituximab, R-CHOP) within last 6 months. More than 69% of VACC-IS patients suffered either severe or moderate COVID-19 disease, which required hospitalisation, whereas 25% of patients were managed supportively at home. All 13 patients had prior SARS-CoV-2 infection with median time of 243 days (IQR: 163-292) prior study enrolment. All patients received second-dose SARS-Cov-2 vaccinations with median time of 115 days (IQR 85-143) prior study enrolment; all patients were double-vaccinated with 7:6 ratio to Pfizer/BNT162b2-to-AZ/ChAdOx1 vaccines. Three patients were recruited who were infection-naïve (HC), with median age of 27 years (IQR: 25-38), and of British-white ethnicity. All HC reported no co-morbidities or on active treatments.

**Table 1.** Patient characteristics of VACC-IS.

| Baseline Characteristics | COVID IS, n=13 |
| --- | --- |
| Age, yr (IQR) | 61 (55-65) |
| Gender |  |
| Male | 7 |
| Female | 6 |
| Ethnicity |  |
| White (%) | 12 (100%) |
| BMI, kg/m<sup>2</sup> (±SEM) | 30.6±2.46 |
| Diagnosis, n (%) |  |
| ESRD requiring transplantation | 7 (53.8%) |
| MPO vasculitis | 1 (7.7%) |
| CLL | 2 (15.3%) |
| Diffuse large B-cell lymphoma | 2 (15.3%) |
| MCD | 1 (7.7%) |
| Co-morbidities, n (%) |  |
| Gout | 2 (15.3%) |
| Hypertension | 6 (46.1%) |
| Type 2 Diabetes Mellitus | 2 (15.3%) |
| DVT | 1 (7.7%) |
| Asthma | 1 (7.7%) |
| Time from positive COVID-19 PCR result, days<br>Median (IQR) | 243 (163-292) |
| WHO COVID-19 clinical severity scale, n % |  |
| Ambulatory mild disease | 4 (30.8%) |
| Hospitalized: moderate disease | 3 (27.7%) |
| Hospitalized: severe disease | 6 (46.1%) |
| Vaccine received, n % |  |
| AZ/ChAdOx1 | 6 (46.1%) |
| Pfizer/BNT162b2 | 7 (58.3%) |
| Time from receiving 2<sup>nd</sup> dose, days<br>Median (IQR) | 115 (85-143) |
| Immunosuppressive regimen, n (%) |  |
| Tacrolimus | 6 (46.1%) |
| Ciclosporin | 1 (7.7%) |
| MMF | 8 (61.5%) |
| Prednisolone | 7 (53.8%) |
| Azathioprine | 1 (7.7%) |
| Rituximab | 3 (23.0%) |
| R-CHOP | 2 (15.3%) |

*ESRD*, End-stage renal disease; *MPO*, Myeloperoxidase; *CLL*, Chronic lymphocytic leukaemia; *ITP*, Immune thrombocytopenia; *MMF*, Mycophenolate Mofetil; R-CHOP, rituximab, cyclophosphamide, doxorubicin hydrochloride, vincristine, and prednisone.

### Cellular response

We examined SARS-CoV-2 cellular response to natural infection and vaccination, by stimulating PBMCs with either Spike (S) (Fig 1a), Nucleocapsid (NC) (Fig 1b), or Membrane (MN) (Fig 1c) peptides, and enumerated IFNγ producing SARS-CoV-2 specific T-cells by ELISpot. As expected, infection-naïve unvaccinated (HC) individuals produced a very low response against S-peptide stimulation, with 2/3 HC eliciting no S-peptide response (Fig 1a). One HC participant demonstrated very low spot forming units (SFU) to S-peptide (3 SFU), which could be a result of non-specific activation. There were statistically significant differences in S-peptide responses between vaccinated immunocompetent (VACC-IC) and immunocompromised (VACC-IS) cohorts versus HC (VACC-IC, p=0.040 and VACC-IS, p=0.025, respectively). No significant differences in S-peptide responses were observed between unvaccinated (UNVACC) versus HC (p=0.471). Comparison of S-peptide responses between VACC-IC and VACC-IS, demonstrated no significant differences (p=0.968, Figure 1a), where, VACC-IS mounted a higher T-cell response to spike (median 17.00 SFU, IQR: 8-44) compared to VACC-IC (median 14.50 SFU, IQR: 7.50-26.75). In VACC-IS, there was one non-responder to S-peptide (0 SFU) which was a kidney transplant patient. Whilst every VACC-IC participant produced a SFU to S-peptide, one participant exhibited 1 SFU, which we categorised as non-responder following two doses of SARS-CoV-2 vaccination. UNVACC had the highest non-responders (n=3), whereby, T-cell responses to S-peptide (median: 5.00, IQR: 1-27) were lower by 9.50 and 12.00 SFU compared to VACC-IC and VACC-IS, respectively.

**Figure 1.**
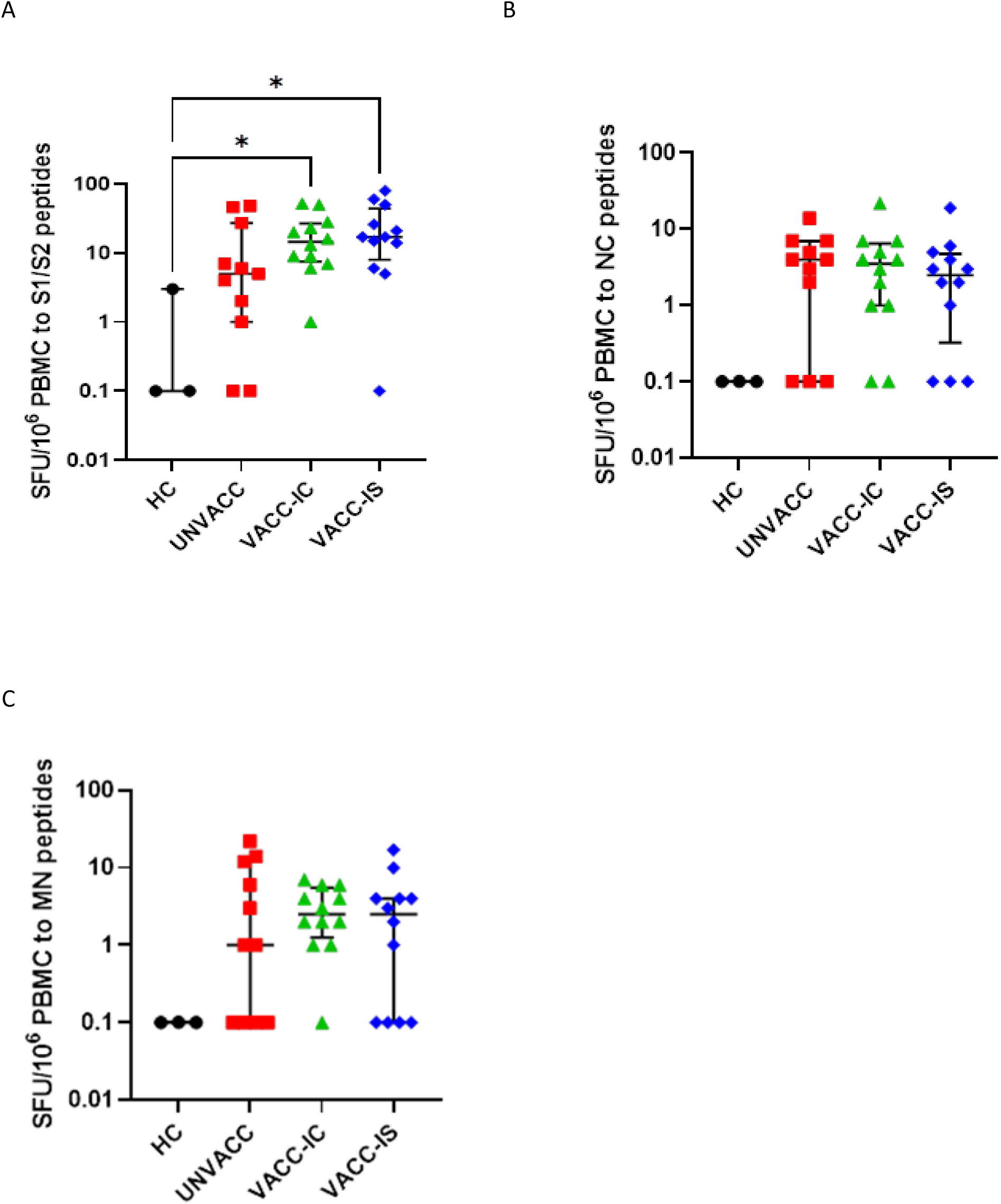
Cellular responses in unvaccinated infection naïve (HC, n=3), unvaccinated convalescent (UNVACC, n=11), vaccinated immunocompetent (VACC-IC, n=12) and vaccinated immunosuppressed (VACC-IS, n=12) participants to Spike (A), Nucleocapsid (B) and Membrane (C) SARS-CoV-2 peptides. One VACC-IS patient was excluded from analysis due to failed positive control. Data representative of individual values expressed as IFNγ spot forming units (SFUs per 2.5×10^6^ PBMCs), median (centre bar) and IQR (upper and lower bars). For visualization of data on log-scale, SFU values=0 is represented by 0.1. Statistical analysis is performed by Kruskal-Wallis nonparametric test with Dunn’s post-hoc test; if not indicated *P* value is not significant, *= *P*<0.05. HC, Healthy infection-naïve; UNVACC, unvaccinated convalescent; VACC-IC, vaccinated immunocompetent; VACC-IS, vaccinated immunosuppressed.

As UNVACC, VACC-IC and VACC-IS cohorts comprised of convalescent individuals, NC and MN responses were detected (Figure 1b, 1c), albeit without any significant differences between all cohorts. Nevertheless, UNVACC and VACC-IS had equal non-responders to NC (n=3) and MN (n=4), whilst non-responders against NC (n=2) and MN (n=1) were also seen in VACC-IC. Moreover, VACC-IC S-peptide responses were significantly higher compared to NC (p=0.005) and MN (p=0.001) by 11.00 and 12.00 SFU, respectively (Figure 2a). Similar trend was observed in VACC-IS (Figure 2b), where S-peptide responses were significantly higher by 14.50 SFU compared to both NC (p=0.005) and MN (p=0.003). Whilst UNVACC S-peptide responses were higher, there were no significant differences between NC and MN (Figure 2c). Together, this demonstrated that both vaccination cohorts induced immunodominance towards S-peptide, whereas no significant precedence to either SARS-CoV-2 peptides were seen with natural infection.

**Figure 2.**
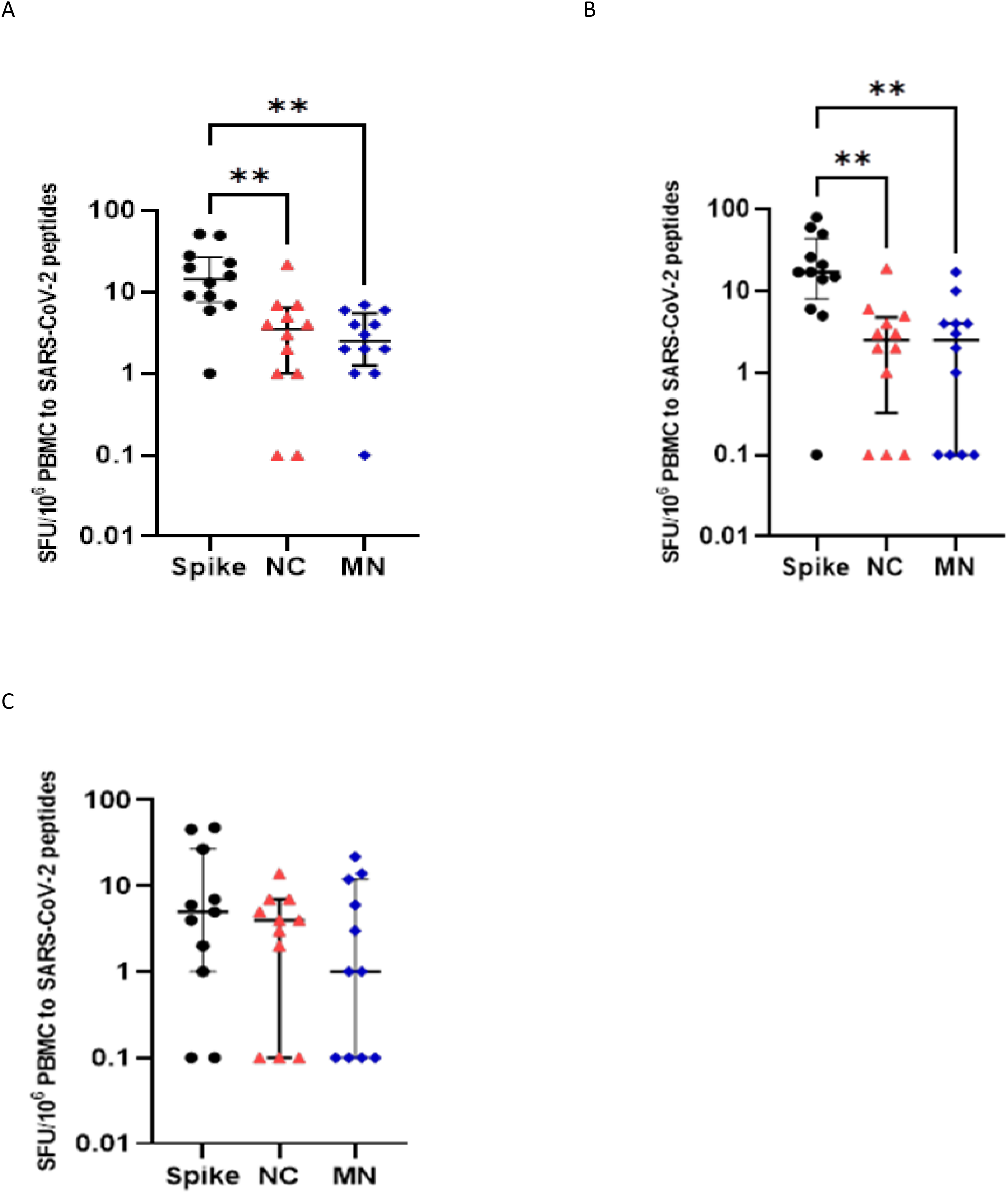
Comparison of Spike, NC, and MN responses in VACC-IC (A), VACC-IS (B) and UNVACC (C) cohorts. One VACC-IS patient was excluded from analysis due to failed positive control. Data representative of individual values expressed as IFNγ spot forming units (SFUs per 2.5×10^6^ PBMCs), median (centre bar) and IQR (upper and lower bars). For visualization of data on log-scale, SFU values=0 is represented by 0.1. Statistical analysis is performed by Kruskal-Wallis nonparametric test with Dunn’s post-hoc test; if not indicated *P* value is not significant, \**P*<0.05, \*\**P*<0.01. HC, Healthy infection-naïve; UNVACC, unvaccinated convalescent; VACC-IC, vaccinated immunocompetent; VACC-IS, vaccinated immunosuppressed.

### Humoral response

Humoral responses were evaluated using total Ig anti-spike ELISA immunoassay. As expected, convalescent unvaccinated and vaccinated cohorts had significantly higher serological responses compared to HC individuals (Fig 3a, p<0.0001). UNVACC cohort demonstrated the lowest serological response (median: 2.92 index-value, IQR: 2.28-3.18), with 10/11 participants displaying seropositivity. Seronegative responses were seen in one UNVACC participant, who also displayed absent SARS-CoV-2 T-cell response. All 12 VACC-IC participants were seropositive (median: 4.40 index-value, IQR: 4.15-4.40), with 10/12 participants generating a serological response which was greater than top standard of assay (4.00 index-value). Moreover, whilst humoral responses were higher in both vaccinated cohorts compared to UNVACC; only VACC-IC demonstrated a significantly higher humoral response compared to UNVACC (p=0.002)

**Figure 3.**
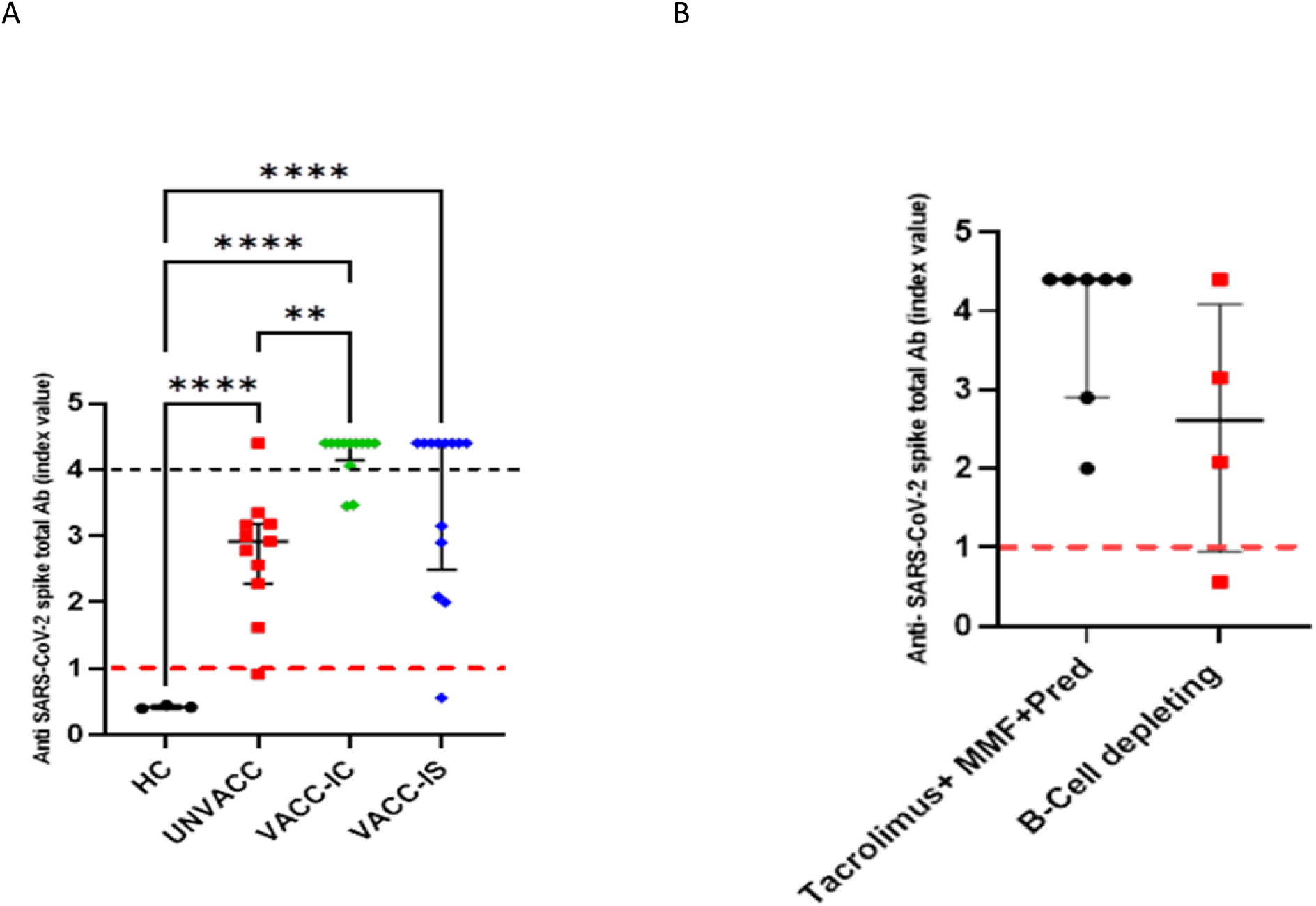
Characterisation of humoral responses in all four cohorts. A.) Anti-SARS-CoV-2 humoral response was evaluated in infection naïve unvaccinated (HC, n=3), convalescent unvaccinated (UNVACC, n=11), convalescent vaccinated immunocompetent (VACC-IC, n=12), and convalescent vaccinated immunocompromised (VACC-IS, n=13) cohorts. Data representative of individual anti-spike SARS CoV-2 total Ab values (index-value), median (centre bar) and IQR (upper and lower bars). B.) Humoral responses in VACC-IS cohort based on immunosuppressive regimes. Responses were categorised into either combinative therapy of Tacrolimus, MMF and Prednisolone versus B-cell depleting therapy (Rituximab and R-CHOP). Median time between patients receiving previous B-cell depleting infusion and serological analysis was 105 days (IQR: 37.50-240.0). Data representative of individual anti-spike SARS CoV-2 total Ab values (index-value), median (centre bar) and IQR (upper and lower bars) for each therapeutic regime. Black dotted lines indicatives of upper limit of assay (4.00 index value). Red dotted lines indicative of assay cut-off threshold for positivity (1.00 index-value). Statistical analysis is performed by Kruskal-Wallis nonparametric test with Dunn’s post-hoc test; if not indicated *p* value is not significant, **p<0.01, ****p<0.0001. MMF, Mycophenolate mofetil; Pred, Prednisolone; R-CHOP, Rituximab, Cyclophosphamide, Doxorubicin Hydrochloride, vincristine, and prednisolone.

In VACC-IS, 12/13 patients were seropositive (median: 4.40 index-value, IQR: 2.49-4.40), with 8/13 patients eliciting responses that were greater than assay top-standard. No significant differences in humoral responses were seen between VACC-IC and VACC-IS. Nevertheless, examining the lower quartile ranges, 25% of VACC-IS participants produced a humoral response (2.49 index-value) which was 1.66 index-value lower than 25% of humoral responses observed in VACC-IC (4.15 index-value). Furthermore, analysing the different immunosuppressive therapies within VACC-IS (Fig 3b), highlighted patients on tacrolimus, MMF and prednisolone, exhibited higher serological response (median: 4.40 index-value, IQR: 2.90-4.40) than those on B-cell depleting therapy (median 2.62 index-value, IQR: 0.94-4.09). Nevertheless, only 1/4 patients receiving B-cell therapy were seronegative. Overall, the above findings highlight both vaccinated cohorts produce higher anti-S serological responses than unvaccinated cohort. Furthermore, vaccinated immunosuppressed patients elicit humoral responses which were comparable to vaccinated immunocompetent cohort.

### Characterisation of CD4^+^ and CD8+ T-cell activation marker expression

To assess phenotypic changes in T-cell activation marker expression post-SARS-CoV-2 peptide stimulation, we performed an unsupervised analysis which evaluates the entire complex scenario depicted by CD4^+^ and CD8^+^ T-cells. Initially, we conducted a dimensionality reduction analysis, flow cytometric and combined t-distribution stochastic neighbour embedding (tSNE), to acquire a phenotypic landscape of CD45^+^CD3^+^CD4^+^ and CD8^+^ lymphocytes in all cohorts (Fig S1). We then explored CD4^+^ and CD8^+^ T-cell panel by unsupervised analysis using FlowSOM [23]. Such analysis conducts multivariate clustering of cells based on self-organised map (SOM) algorithm, enabling cells to be stratified into specific meta-clusters based on HLA-DR and CD38 expression [24]. Heat maps were used to statistically report differences in phenotypic expression between cohorts.

For CD4^+^ characterisation (Fig 4a-b) against S-peptide stimulation, we clustered all individual cells for each cohort into 15 distinct clusters based on surface HLA-DR and CD38 expression. Subsequently, we reduced complexity by merging similar cluster profiles and conducted further re-clustering. As illustrated in Fig 4a, 5 distinct clusters in CD4^+^ T-cells were identified in all cohorts. Each metacluster were represented equally within all cohorts, with exception to metacluster 4, which was lower in VACC-IS. Moreover, no significant differences in CD4^+^HLA-DR CD38 phenotypes were seen between cohorts (Fig 4b). Dual HLA-DR^+^CD38^+^ expression was identified in metacluster 4 and 9, whilst metacluster 7 illustrated CD4^+^ HLA-DR^+^CD38^wk^ expression. Both metacluster 3 and 10 portrayed HLA-DR^−^CD38^+^ and HLA-DR^+^CD38^−^, respectively. Overall CD4^+^ HLA-DR^+^ expression was in 80% of metaclusters (n=4), whilst CD38^+^, CD38^−^ and CD38^wk^, were identified in 60% (n=3), 20% (n=1) and 20% (n=1), respectively.

**Figure 4.**
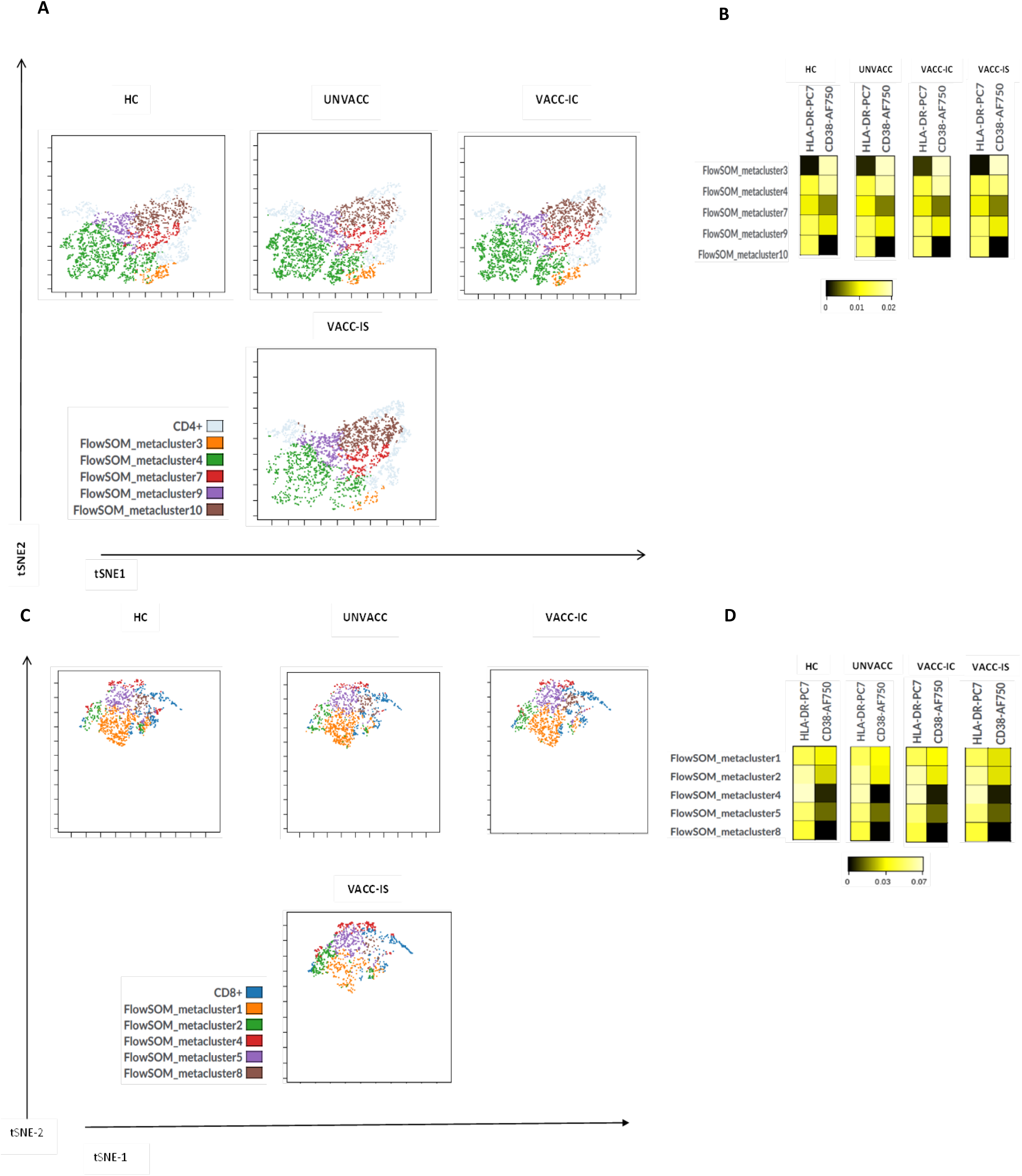
Unsupervised analysis of CD4^+^ and CD8^+^ T-cells post S-peptide stimulation. **A** Representation of CD4+ phenotypic landscape, by coupling tSNE dimensional-reductional analysis with FlowSOM which was used to identify specific CD4+ T-cell metaclusters based on HLA-DR and CD38 expression for each cohort. **B** Heat map representing the different CD4+ metaclusters identified by FlowSOM for each cohort, where the colours in the heatmap represent the median acrsinh ratio for HLA-DR and CD38 expression of each metacluster. Heatmap colours vary from black for lower expression, to yellow for higher expression of each surface marker (HLA-DR, CD38). **C** The same unsupervised analysis was used to define the CD8+ phenotypic landscape, coupled with FlowSOM, for identification of CD8+ metaclusters between cohorts. **D** Heat map representing the median arcsinh ratio of HLA-DR and CD38 observed for each cohort.

Characterisation of CD8^+^ S-peptide responses demonstrated 5 clusters (Fig 4c), where metacluster 1 and 8 were under-represented in VACC-IS cohort, whilst remaining metaclusters were similar across cohorts. Unlike CD4^+^, there were differences in HLA-DR and CD38 expression observed between cohorts, as illustrated by metacluster 1 and 2 (Fig 4d). CD8^+^ HLA-DR^+^CD38^wk^ expression were identified in VACC-IS metacluster 1 (as represented by darker yellow shade for CD38), whereas dual HLA-DR^+^CD38^+^ expression were seen in HC, UNVACC and VACC-IC. Moreover, dual HLA-DR^+^CD38^+^ expression in UNVACC and VACC-IC were identified for metacluster 2, whereas significant different outcomes were observed in HC and VACC-IS; as illustrated by HLA-DR^+^CD38^wk^ expression. Furthermore, all CD8^+^ metaclusters expressed HLA-DR^+^(n=5), whereas CD38^+^ were reported in 20% and 40% of metaclusters in HC (n=1) and UNVACC, VACC-IC (both n=2), respectively. No CD38^+^ expression were identified in VACC-IS, with 60% of VACC-IS metaclusters (n=3) expressing CD38^wk^, and 40% (n=2) were CD38^−^. Overall post S-peptide stimulation, both CD4^+^ and CD8^+^ upregulated HLA-DR^+^, with higher CD38^+^ observed in CD4^+^ T-cells.

Evaluation of HLA-DR and CD38 expression was conducted in NC (Fig S2a-d). 7 metaclusters were identified for NC. As with S-peptide, under-representation was observed in VACC-IS (metacluster 6, Fig S2a), which expressed HLA-DR^wk^CD38^+^ (Fig S2b). Moreover, dual CD4^+^ HLA-DR^+^ CD38^+^ were seen in metacluster 4 in all cohorts (Fig S2b); albeit expression of HLA-DR and CD38 was lower in both vaccinated cohorts compared to UNVACC and HC. Overall, NC-peptide stimulation preferentially expressed CD38^+^ (n=4) compared to HLA-DR^+^ (n=2) in CD4^+^. For CD8^+^ NC responses, 4 metaclusters were identified (Fig S2c), metacluster 7 and 2 were under-represented in VACC-IS and UNVACC, respectively. Both metacluster 7 and 2 expressed CD8^+^HLA-DR^+^CD38^+^ and CD8^+^HLA-DR^+^CD38^−^, respectively. (Fig S2d). Furthermore, CD8^+^ stimulated with NC-peptide elicited a predominant dual CD8^+^ HLA-DR^+^ CD38^+^ phenotype (n=3) in all cohorts, with only metacluster 2 demonstrating a CD8^+^ HLA-DR^+^ CD38^−^ phenotype. Overall, a greater T-cell activation profile was observed in CD8^+^, compared to CD4^+^, following NC-peptide stimulation.

Stimulation with MN-peptide resulted in characterisation of 7 CD4^+^ metaclusters (Fig S3a). The similar trend of metacluster under-representation was evident in VACC-IS CD4^+^ (metacluster 1), which depicted an HLA-DR^wk^CD38^+^ profile (Fig S3b). No dual CD4^+^ HLA-DR^+^ CD38^+^ were seen across all cohorts, with HLA-DR^wk^ expression observed across all metaclusters, whilst 57%(n=4) of CD4^+^ metaclusters expressed CD38^wk^; only 3 metaclusters depicted CD38^+^. However, within CD8^+^ landscape, UNVACC cohort depicted a substantial drop in metacluster 5 and 9 representation (Fig S3c) with HLA-DR^+^ CD38^−^ and HLA-DR^+^ CD38^+^ expression (Fig S3d), respectively. Dual HLA-DR^+^CD38^+^ expression was observed in metacluster 9, with HLA-DR^+^ expressed in all CD8^+^ metaclusters (n=5); albeit at lower levels in metacluster 14. CD38^+^ expression was generally lower in CD8^+^ metaclusters across all cohorts, with VACC-IS and HC illustrating the lowest CD8^+^CD38^+^ expression. Overall, in S, NC and MN-peptide stimulations, VACC-IS exhibited lower proportions of specific metaclusters. Furthermore, HLA-DR^+^ was preferentially upregulated in CD8^+^ whereas, CD38^+^ expression was skewed towards CD4^+^ T-cells.

### Characterisation of T-cell subsets

As proof-of-principle, we conducted a T-cell subset (TCS) immunophenotyping panel, post-S-peptide stimulation, on VACC-IC (n=3) and VACC-IS (n=2) participants. Initially, we used a manual gating strategy (Fig S4) where we compared both CD4^+^ and CD8^+^ T-cell subsets as illustrated in Figure 5a-b. Within CD4^+^ and CD8^+^ populations, we examined markers for T-cell differentiation (CD45RA, CD197, CD27 and CD28), senescence and exhaustion (CD57 and CD279 (PD1), respectively). As highlighted in Fig 5a, there were no significant differences between CD4^+^ T-cell subsets between VACC-IC and VACC-IS. Both CD4^+^ naïve (T_n_) and effector (T_e_) cells were higher, by 10.29% and 6.39%, respectively, in VACC-IC compared to VACC-IS. Whereas CD4^+^ T-effector-memory (T_em_) was higher by 16.74% in VACC-IS compared to VACC-IC. Both VACC-IC and VACC-IS had very similar T-central memory (T_cm_) populations. Furthermore, senescent CD4^+^ T-cells (CD4^+^CD57^+^) were also similar between both cohorts; except for one VACC-IC participant who exhibited higher CD4^+^CD57^+^ cells (19.62%, Fig 5a).

**Figure 5.**
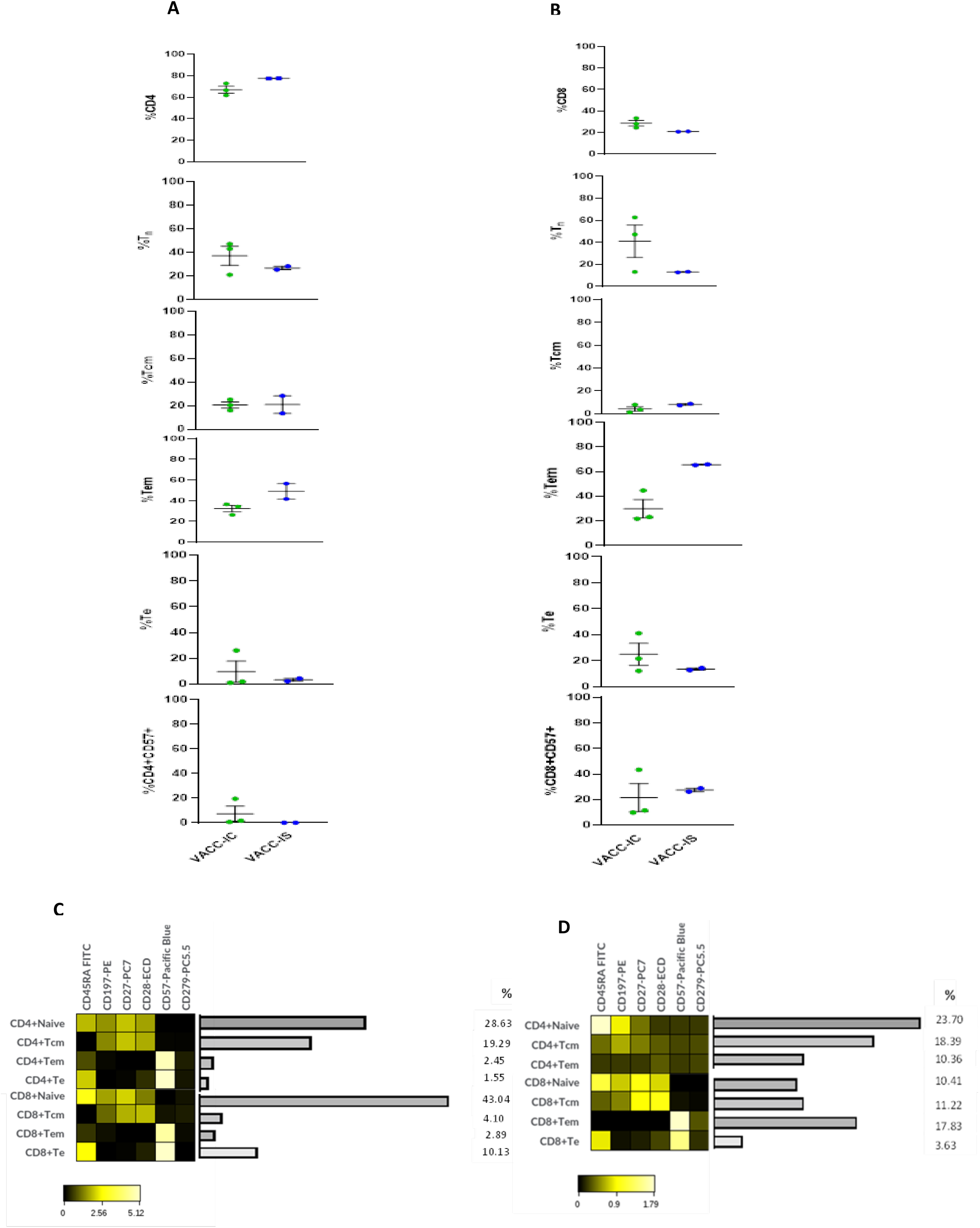
Characterisation of T-cell subsets. Percentages of different CD4^+^ T-cell (**A**) and CD8^+^ T-cell subpopulations (B) are shown for VACC-IC (n=3) and VACC-IS (n=2), as obtained by manual gating strategy. Data representative of individual values, mean (centre bar) ±SEM (upper and lower bars). Statistical analysis conducted using two-sided Mann-Whitney test; if not indicated, p-value not significant. **C**. Heat map representing different CD4^+^ and CD8^+^ T-cell subpopulations for VACC-IC (C) and VACC-IS (**D**) cohorts, as identified by tiSNE. The tSNE plot was designed by concatenation of samples per cohort where equal sampling of 60,845 and 16,388 for CD4^+^ and CD8^+^, respectively, were used for VACC-IC. For VACC-IS, 60,932 CD4^+^ and 16,397 CD8^+^ events were used. The colours in the heat map represents the median acrsinh ratio for each surface marker expression. Heatmap colours vary from black for lower expression, to yellow for higher expression. Tn is identified as CD45RA^+^CD197^+^CD27^+^CD28^+^; Tcm are CD45RA^−^CD197^+^CD28^+^CD27^+/−^; Tem are CD45RA^−^CD197^−^CD28^−^CD27^+/−^; Te are CD45RA^+^CD197^−^CD28^−^CD27^−^CD57^+^; Exhausted cells express CD57^+^CD279^+^ (PD1). Tn, Naïve T-cells; Tcm, T-central memory; Tem, T-effector memory; Te, T-effector.

A similar trend was seen in CD8^+^ subsets, where T_n_ and T_e_ were higher in VACC-IC, whilst T_em_ populations higher in VACC-IS (Figure 5b). Moreover, both VACC-IC and VACC-IS demonstrated a 15.19% and 10.14% increase in CD8^+^ T_e_, respectively, compared to CD4^+^ T_e_. These observations were supported with elevated senescent-terminally differentiated CD8^+^ (CD8^+^CD57^+^) levels compared to CD4^+^. Overall, with exception of T_n_, CD4^+^ and CD8^+^ T_em,_ displayed highest percentage values across both cohorts. Subsequently, Tem was identified as the predominant memory T-cell subset. Furthermore, no exhausted CD4^+^ and 8^+^ T-cells (CD4^+^CD57^+^PD1^+^) were seen in both cohorts using manual gating strategy.

Subsequently, we used the same unsupervised analysis, as used for activation-markers, where tiSNE analysis identified CD4^+^ and CD8^+^ TCS, whose percentages are represented alongside the heatmap; which portrays TCS marker expression in both cohorts (Fig 5c-d). Immediately, it can be recognised VACC-IS had absent CD4^+^ T_e_ subset (Fig 5d), whereas a low percentage (1.55%) were identified for VACC-IC (Fig 5c). Unlike manual gating strategy, tSNE analysis demonstrated CD4+T_cm_ were dominant for VACC-IC (19.29%) and VACC-IS (18.39%); which were similar between both cohorts. Whereas, VACC-IS evoked a 7.91% increase in CD4+T_em_ compared to VACC-IC. Furthermore, as highlighted in Figure 5d, VACC-IS CD4^+^ T_n_ exhibited considerably weaker CD27 and CD28 expression, compared to VACC-IC. Furthermore, VACC-IS CD4^+^ T_n_, along with T_cm_, T_em_ and CD8^+^ T_em_ exhibited weak expression of exhausted T-cells (CD57 PD1, dark green on heatmap); whereas such CD57 PD1 phenotypes were absent in these subsets in VACC-IC. These subtle variations were not detected from use of manual gating strategy.

Both VACC-IS CD8^+^ T_cm_ and T_em_ were greater by 7.12% and 14.94% (Fig 5d), respectively, compared to VACC-IC (Fig 5c). Whereas VACC-IC depicted a 6.50% increase in CD8+ T_e_ compared to VACC-IS. Overall, tSNE analysis demonstrated CD4^+^T_cm_ as the dominant memory T-cell subset in both VACC-IC and VACC-IS. Whereas CD8^+^ T_e_ and T_em_ were the dominant subset in VACC-IC and VACC-IS, respectively, post S-peptide stimulation.

### *Ex vivo* production of pro-inflammatory cytokines

Multiplex cytokine analysis (IL-6, TNFα, IL-1β and IL-10) was performed on study cohorts after antigen-specific stimulation in PBMCs with SARS-CoV-2 S, NC, and MN peptides (Fig 6a-c). We recognise IFNγ secretion as a key cytokine signature in viral infections [25], however as PBMCs were harvested within an IFNγ capture ELISpot plate this cytokine was excluded.

**Figure 6.**
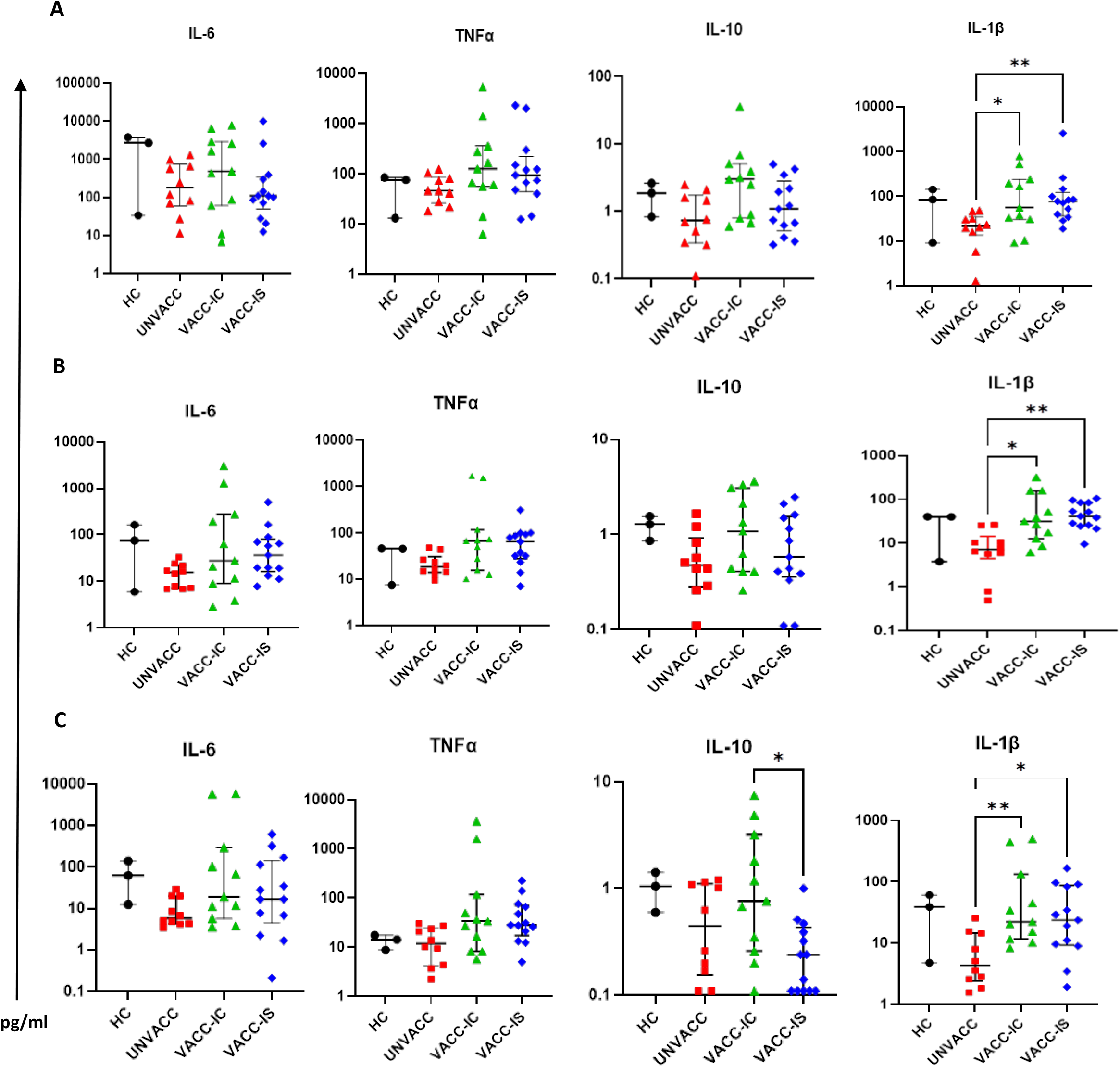
Cytokine secretion following SARS-CoV-2 peptide stimulation. Multiplex cytokine analysis was conducted using supernatant after antigenic-specific stimulation with Spike (A), Nucleocapsid (NC), and Membrane (MN) peptides in HC (n=3), UNVACC (n=10), VACC-IC (n=11) and VACC-IS (n=13) participants. Two samples from UNVACC (n=1) and VACC-IC (n=1) were excluded due to laboratory technical error. Individual data points are illustrated as individual scatter plots for each cytokine, expressed as pg/ml, with median (centre bar) and IQR (upper and lower bars). Statistical analyses were determined using nonparametric Kruskal-Wallis test with Dunn’s post-hoc test for multiple comparisons. \**P*<0.05, \*\**P*<0.01. HC, Healthy infection-naïve; UNVACC, unvaccinated convalescent; VACC-IC, vaccinated immunocompetent; VACC-IS, vaccinated immunosuppressed.

Of the four cytokines analysed, three (IL-6, TNFα and IL-10) showed no significant differences in secretion between cohorts following S-peptide stimulation (Fig 6a). IL-1β levels were significantly elevated in both VACC-IC (median 55.85 pg/ml IQR: 30.58-241.4) and VACC-IS (median 77.25 pg/ml, IQR: 30.58-241.4) compared to UNVACC (Fig 6d, p=0.023 and p=0.008, respectively). IL-6 secretion demonstrated the highest magnitude of cytokine secretion in all cohorts after S-peptide stimulation. Interestingly, HC participants secreted the highest IL-6 levels (median 2711 pg/ml, IQR: 33.61-3779), whereas within convalescent cohorts, VACC-IC produced the highest IL-6 levels (median 482.4 pg/ml, IQR: 61.16-2894), which was 300.7and 372.7 pg/ml greater than UNVACC and VACC-IS, respectively. Similarly, VACC-IC secreted the highest TNFα levels (median 126.7 pg/ml, IQR: 55.97-362.5) which were 50.6, 80.47, and 31.29 pg/ml higher than HC, UNVACC and VACC-IS.

Following NC-peptide stimulation, no significant differences in IL-6, TNFα and IL-10, were observed between cohorts (Fig 6b). Similar to S-peptide, significantly elevated IL-1β levels were detected in VACC-IC and VACC-IS compared to UNVACC in both NC (p=0.023 and p=0.004, respectively) and MN-peptide stimulation (p=0.005 and p=0.025, respectively, Fig 6c). Furthermore, following MN-peptide stimulation (Fig 6c), IL-10 levels were modestly elevated in VACC-IC compared to VACC-IS (p=0.033). Overall, the magnitude of cytokine secretion observed across convalescent cohorts (UNVACC, VACC-IC and VACC-IS) were all significantly greater for IL-6, TNFα and IL-1β than IL-10 (Fig S5). These findings are indicative of a strong bias towards Th1 cytokine secretion following S, NC and MN-peptide stimulation.

## DISCUSSION

Immunisation represents the most effective intervention against infectious diseases, such as SARS-CoV-2; as evident by success of mass global vaccination programmes reducing viral spread and preventing severe disease [26]. Nevertheless, there are very few studies exploring the immunogenicity of COVID-19 vaccines in immunocompromised patients, such as solid-organ transplant recipients (SOTs) and haematological malignancies. Moreover, reduced vaccine-induced immune responses have been associated in SOTs, or in general, in patients on active immunosuppressive therapies [27]. To address this, we explored the immunogenicity of two SARS-CoV-2-19 licenced vaccines (either BNT162b2 mRNA or ChAdOx1 nCoV-19 adenoviral-vector) in double-vaccinated adult renal transplant recipients and those diagnosed with haematological malignancies. Unlike previous studies where vaccine immunogenicity was limited to early post-vaccine period [28], we enrolled immunosuppressed patients with a median time of 115 days post second-dose, thus, providing an up-to-date snapshot of their immune response to SARS-CoV-2 vaccination. We compared the humoral and cellular responses of this immunocompromised group (VACC-IS) to healthy vaccinated (VACC-IC), unvaccinated (UNVACC) and infection-naïve (HC) cohorts. Our data demonstrates that VACC-IS patients responded to the vaccine by producing comparable cellular and humoral responses to VACC-IC. However, findings from large prospective studies [29] are required to correlate such vaccine-induced response with protective immunity.

Recent reports have highlighted diminished T-cell responses against COVID-19 vaccines in renal transplants patients receiving T-cell directed therapies [28] and in haematological cancer patients [30]. In response to these studies we examined vaccine-induced SARS-CoV-2 specific T-cell responses in these patients through using an IFNγ release assay. Reassuringly, 92% of VACC-IS patients (n=12) elicited a detectable T-cell response following spike (S) peptide stimulation. An identical T-cell response rate were observed in VACC-IC participants, demonstrating no differences in vaccine-induced T-cell responses between VACC-IC and VACC-IS. Interestingly, a 17.24 % increase in IFNγ-secreting SARS-CoV-2 specific T-cells were identified in VACC-IS compared to VACC-IC. Our findings are consistent with the preliminary OCTAVE trial data (ISRCTN: 12821688), where T-cell responses were similar across immunosuppressed and immunocompetent cohorts [29]. Similarly, both VACC-IC and VACC-IS PBMCs stimulated with SARS-CoV-2 peptides induced a predominantly Th1 response, with significantly elevated IL-6, TNFα and IL-1β compared to IL-10. Moreover, cytokine and T-cell responses in vaccinated cohorts demonstrated immunodominance towards S-peptide compared to NC an MN peptides; findings that are consistent with both vaccine clinical trials [19,20]

Both VACC-IC and VACC-IS had one T-cell non-responder each; however, both these participants demonstrated positive serology for anti-spike SARS-CoV-2 antibodies. Moreover, 3 VACC-IS patients had received B-cell depleting therapy (Rituximab) 2 months following their second SARS-CoV-2 vaccine dose. Whilst 2/3 of these patients, were serologically positive, their anti-spike antibody levels were lower in comparison to those receiving T-cell targeted therapies. Such correlation between diminished vaccine specific humoral responses and B-cell depleting therapies have been reported in prior studies [31]. Nevertheless, all 3 patients receiving B-cell therapy, including the patient who failed to seroconvert, elicited T-cell responses to S-peptide stimulation. These results highlight that B-cell negative patients, due to primary or therapy-induced aetiologies, can still reap benefit from T-cell compartment of vaccination.

A plausible explanation for comparable T-cell responses observed in our data, which were not seen in a study conducted by Prendecki et al [28], could be that our immunosuppressed patients had prior natural infection, subsequently it could represent an augmented response to second-dose vaccine (“third” challenge in these convalescent patients). In fact, the same study reported a 54% increase in T-cell response in their immunosuppressed patients following second-dose vaccination [28]. Such findings support additional vaccine doses could provide an immunogenic “top-up” in immunosuppressed patients. Going forward, we propose a comparative evaluation of assessing vaccine immunogenicity between convalescent and infection-naïve vaccinated immunosuppressed patients. Findings from these studies could provide evidence-based data for optimal vaccine type and dosing schedule in these patients.

All study participants, except HC-infection naïve, had prior natural infection, where unvaccinated (UNVACC) had the highest T-cell non-responders (n=3) to S-peptide stimulation. Interestingly, one UNVACC T-cell non-responder was also seronegative for anti-spike antibodies. This participant represented a house-hold case of COVID-19; with positive real-time polymerase chain-reaction nasopharyngeal result, and no significant medical history. We speculate one of two reasonings; firstly, this could represent a case of natural waning immunity, or, secondly, a false-positive result. We believe the latter is unlikely, as the house-hold contact was tested in our study and had detectable serology and T-cell response. Furthermore, all but one HC participant had no detectable T-cell responses. One HC participant had a weak response of 3 SFU to S-peptide. We favour two hypothetical models which could explain this. Firstly, this result could represent a cross-reaction with other six human pathogenic coronaviruses [32]. Secondly, as these were healthcare workers, both occupational and house-hold exposure could evoke very low concentration of SARS-CoV-2, which may be insufficient to elicit a B-cell response but may induce a T-cell response.

Investigating the CD4^+^ and CD8^+^ vaccine-induced landscape highlighted key differences between VACC-IC and VACC-IS. Firstly, whilst no significant differences in CD4^+^ surface activation markers (CD38 and HLA-DR) were observed between VACC-IC and VACC-IS, the abundance of the dominant metacluster population were reduced in VACC-IS. Similarly, reduction of metacluster abundance were identified in VACC-IS CD8^+^, however, with notable differences in T-cell activation marker expression. Over 40% of VACC-IC CD8^+^ metaclusters depicted dual HLA-DR^+^CD38^+^ expression with elevated levels of CD8^+^ T_e_ (CD45RA^+^CD197^−^CD27^−^CD28^−^) cells post-S-peptide stimulation. Such finding is consistent with prior studies which have highlighted terminal effector T-cells overexpress the activation markers CD38 and HLA-DR [33]. However, the same metaclusters were identified as HLA-DR^+^CD38^wk^ in VACC-IS. Moreover, VACC-IS demonstrated a greater increase in CD8^+^ T_em_ (CD45RA^−^CD197^−^CD27^+/−^CD28^−^ CD57^+^) subsets compared to VACC-IC. Such findings may explain the increased levels of IFNγ secreting SARS-CoV-2 specific T-cells observed in our ELISpot data; as CD8+T_em_ have shown to secrete the greatest IFNγ levels compared to other T-cell memory subsets [34].

Similar findings have been reported in a recent study investigating vaccine-induced response in multiple sclerosis patients on anti-CD20 therapy [35]. No differences in T-cell activation were seen in CD4^+^ compartments post-vaccination in both healthy and MS-patient cohorts. However, CD8^+^ HLA-DR^+^CD38^wk^ metaclusters were seen in MS patients, which were predominantly of the T_em_ subset in line with our findings in VACC-IS cohort. The authors concluded such findings in MS-patients are indicative of a robust CD8^+^ T-cell response compared to healthy controls. However, we hypothesise the lack of CD4^+^ T-follicular helper cells and vaccine-induced antibodies could have preferentially driven and augmented CD8^+^ T-cell responses. Whilst these findings are encouraging, we believe extensive deep-immune profiling comprising a broader range of immunosuppressed patients are required to achieve a definitive illustration of vaccine-induced T-cell responses. We propose undertaking activation-induced marker (AIM) assays on CD4^+^ and CD8^+^ antigenic specific cells. Such experimental design may provide in-depth information surrounding CD4^+^ T-cell priming by examining co-expression of CD200 and CD40L. Functional CD8^+^ T-cell responses can be investigated through examining IFNγ, TNFα, IL-2 and granzyme B expression. Such functionality could be correlated with polyfunctional status of SARS-CoV-2 specific T-cells as it remains unclear whether mono-or polyfunctional T-cells are of greater protective value [19].

Our study has some limitations. Firstly, the small number of participants and immunosuppressed patients, restricted to renal transplant and haematological malignancies, makes it challenging to draw firm conclusions. Moreover, demographical risk factors for COVID-19, such as ethnicity [36] were not controlled for as most participants were white-British. Consequently, these demographic variables could not be fully investigated in this study. Secondly, we were only able to re-bleed a small proportion of our VACC-IC and VACC-IS study participants for T-cell subset analysis. Going forward, we propose to extend the T-cell subset panel along with drop-in markers of activation and proliferation (such as Ki-67). This would provide a more detailed phenotypic landscape of T-cell memory subsets found in vaccinated healthy and immunosuppressed cohorts.

Overall, our data confirms an immunological response to SARS-CoV-2 vaccines in immunosuppressed patients, when assessed by combination of cellular and serological assays. The observed vaccine-induced responses within this immunosuppressed cohort were comparable to healthy vaccinated participants. Furthermore, our data highlights the robust and broad capacities of SARS-CoV-2 specific T-cells. Further work is required to decipher these responses with the continual emergence of global SARS-CoV-2 variants of concerns. Our findings warrant further work correlating the observed immunological responses with protective immunity and evaluate if longevity of these responses is comparable to healthy individuals. Such information may aid development of a standardised immunisation schedule required to optimise the vaccine-induced responses observed in this clinically vulnerable patient group.

## Data Availability

Data available upon request

## Author Contributions

Conceptualisation, D.M and A.W.; Methodology, D.M, A.W and S.S; Funding Acquisition, D.M, A.W, M.M; Investigation, S.B, D.M, A.G, S.S; Writing-review and editing, D.M, A.W

## Institutional Review Board Statement

Study proposal was evaluated and approved by Portsmouth NHS Trust Research Ethics Committee (Study reference: 21/BPR10029) and national Health Research Authority ethics committee (London-City and East research ethics committee, IRAS: 291009).

## Informed Consent Statement

Informed consent was obtained from all participants involved in this study.

## Data Availability Statement

Data available upon request

## Acknowledgments

We would like to thank Lynn Watkins, Lynn Vinall and Marie Broadway (Renal research team, Portsmouth Hospital NHS University Trust) for retrieving blood samples from our study participants. We would also like to thank all participants and patients involved in this study.

