## Supplemental figures and tables for "Cellular and humoral responses to SARS-CoV-2 vaccination in immunosuppressed patients"

| Baseline Characteristics | UNVACC n=11 |
| --- | --- |
| Age, yr (IQR) | 37 (31-47) |
| Gender  Male  Female  Ethnicity  White (%)  Black  Asian  BMI, kg/m^2^ (±SEM)  Co-morbidities, n (%)  Hypothyroidism  Stroke  Sleep Apnea  Time from positive COVID-19 PCR result, days  Median (IQR)  WHO COVID-19 clinical severity scale, n %  Ambulatory mild disease  Hospitalized: moderate disease  Occupation, n (%)  Patient-facing  Non-patient facing | 3  8  9 (81.8%)  1 (9%)  1 (9%)  33.9±1.9  1 (9%)  1 (9%)  2 (18.1%)  160 (145-165)  9 (81.8%)  2 (18.1%)  7 (63.6%)  4 (36.4%) |

**Supplemental table S1. Patient characteristics of UNVACC.**

| Baseline Characteristics | VACC-IC, n=12 |
| --- | --- |
| Age, yr (IQR) | 45 (30-53) |
| Gender  Male  Female  Ethnicity  White (%)  Asian (%)  BMI, kg/m^2^ (±SEM)  Co-morbidities, n (%)  Depression  Asthma-controlled  Time from positive COVID-19 PCR result, days  Median (IQR)  WHO COVID-19 clinical severity scale, n %  Ambulatory mild disease  Vaccine received, n %  Pfizer/BNT162b2  Time from receiving 2^nd^ dose, days  Median (IQR)  Occupation, n (%)  Patient-facing  Non-patient facing | 2  10  10 (83%)  1 (17%)  24.2±1.48  2 (22.2%)  3 (11.1%)  175 (143-431)  11 (100%)  11 (100%)  112.5 (87.2-153)  6 (50%)  6 (50%) |

**Supplemental table S2. Patient characteristics of VACC-IC.**

**Figure S1**

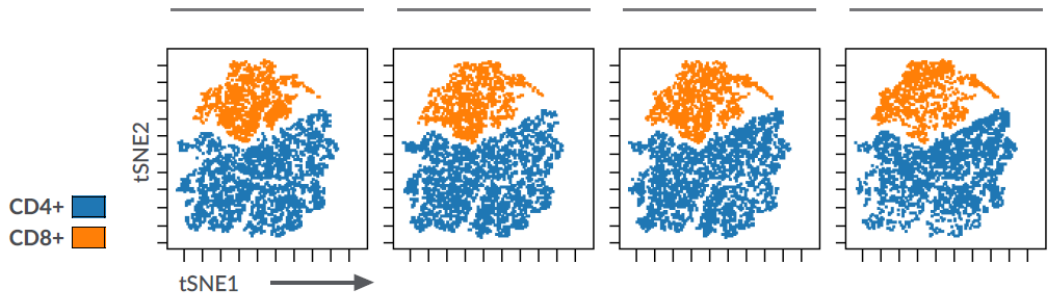

HC

UNVACC

VACC-IC

VACC-IS

**Figure S1. Immunophenotyping of T-lymphocyte populations post SARS-CoV-2 peptide stimulation**. tSNE analysis of 103,311 CD45^+^ CD3^+^ lymphocytes from 37 study participants across four cohorts. One sample from VACC-IC and VACC-IS (n=2) were excluded from analysis due to low event count. The tSNE plot was designed by concatenation of samples per cohort, where equal sampling of 4453 events was selected from each cohort.

**Figure S2**

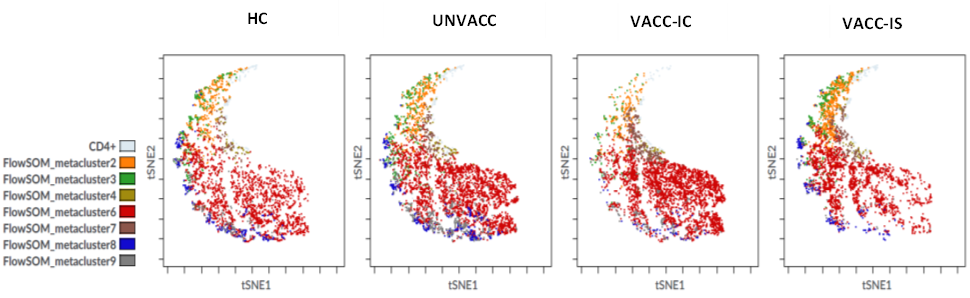

**A**

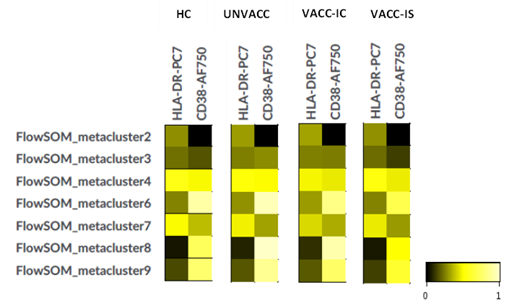

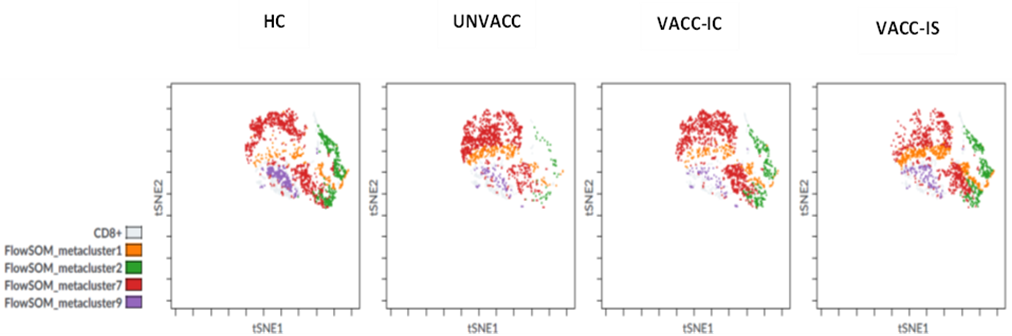

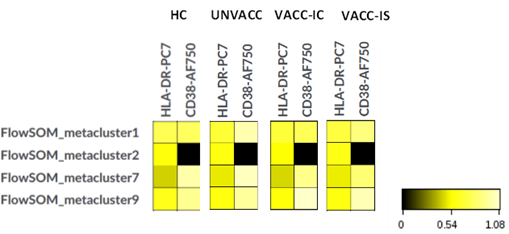

**B**

**C**

**D**

**Figure S2. Unsupervised analysis of CD4^+^ and CD8^+^ T-cells post NC-peptide stimulation.** **A** Representation of CD4^+^ phenotypic landscape, by coupling tSNE dimensional-reductional analysis with FlowSOM which was used to identify specific CD4^+^ T-cell metaclusters based on HLA-DR and CD38 expression for each cohort. **B** Heat map representing the different CD4^+^ metaclusters identified by FlowSOM for each cohort, where the colours in the heatmap represent the median acrsinh ratio for HLA-DR and CD38 expression of each metacluster. Heatmap colours vary from black for lower expression, to yellow for higher expression of each surface marker (HLA-DR, CD38). **C** The same unsupervised analysis was used to define the CD8^+^ phenotypic landscape, coupled with FlowSOM, for identification of CD8^+^ metaclusters between cohorts. **D** Heat map representing the median arcsinh ratio of HLA-DR and CD38 observed for each cohort.

**Figure S3**

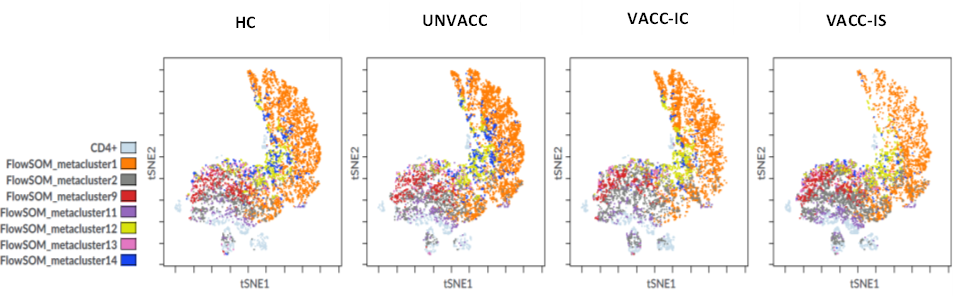

**A**

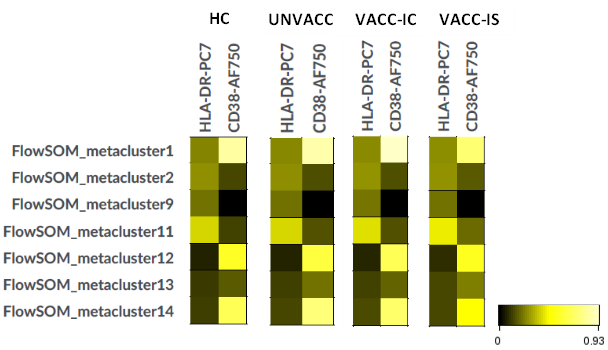

**B**

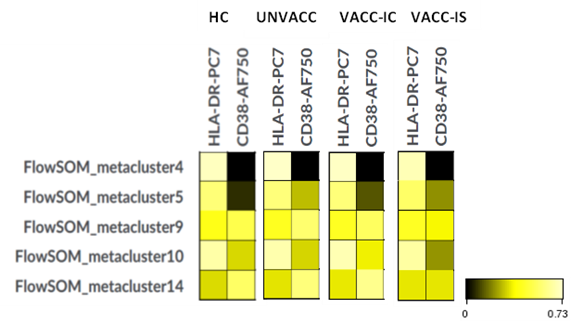

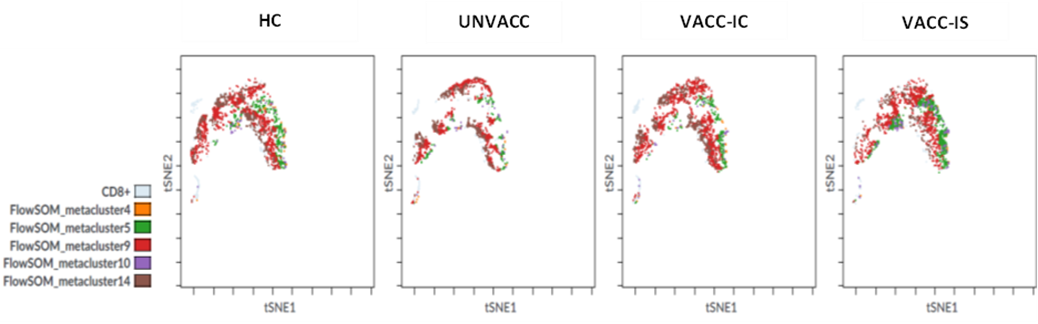

**C**

**D**

**Figure S3. Unsupervised analysis of CD4^+^ and CD8^+^ T-cells post MN-peptide stimulation.** **A** Representation of CD4^+^ phenotypic landscape, by coupling tSNE dimensional-reductional analysis with FlowSOM which was used to identify specific CD4^+^ T-cell metaclusters based on HLA-DR and CD38 expression for each cohort. **B** Heat map representing the different CD4^+^ metaclusters identified by FlowSOM for each cohort, where the colours in the heatmap represent the median acrsinh ratio for HLA-DR and CD38 expression of each metacluster. Heatmap colours vary from black for lower expression, to yellow for higher expression of each surface marker (HLA-DR, CD38). **C** The same unsupervised analysis was used to define the CD8^+^ phenotypic landscape, coupled with FlowSOM, for identification of CD8^+^ metaclusters between cohorts. **D** Heat map representing the median arcsinh ratio of HLA-DR and CD38 observed for each cohort.

**Figure S4**

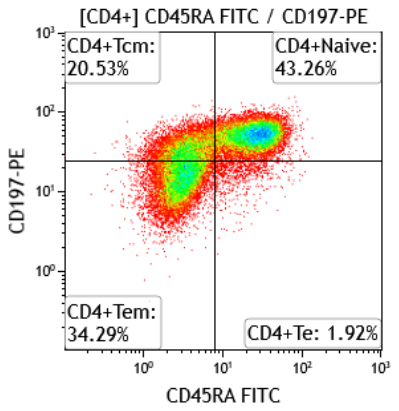

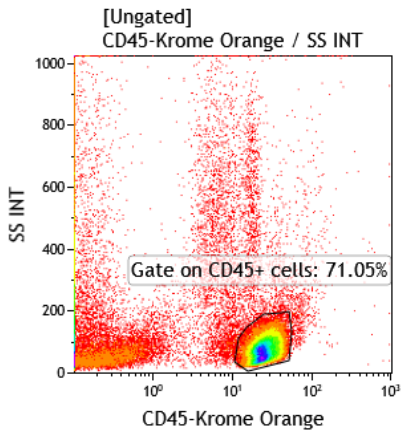

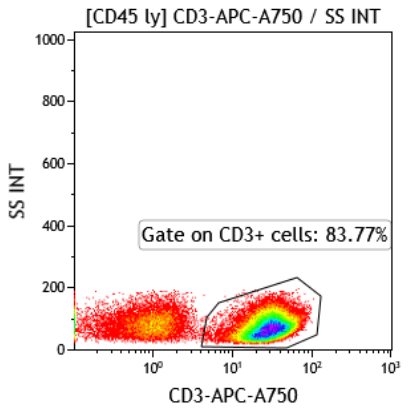

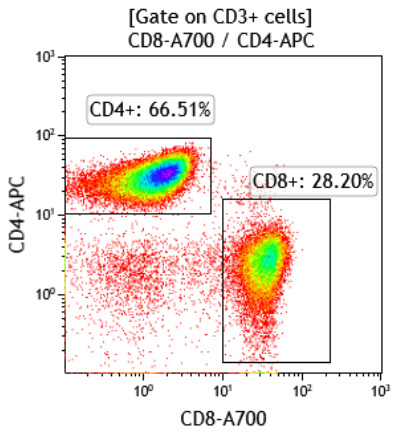

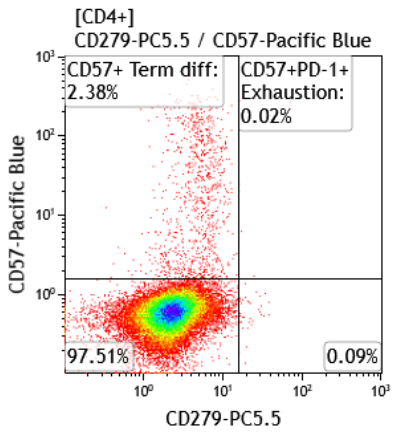

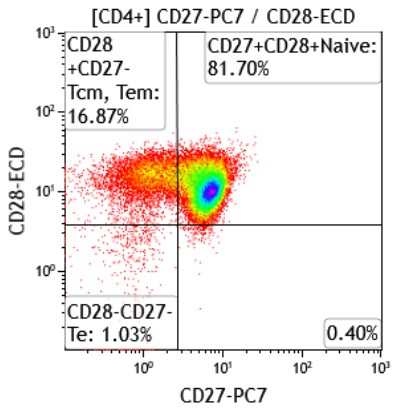

**Differentiation**

**Senescence/Exhaustion**

**Figure S4. Gating strategy used for characterisation of CD4^+^ and CD8^+^ T-cell subsets.** The first CD45+ gate was used to identify the lymphocyte population. This was then sequentially gated on CD3+ T-cells, which was required to identify population of interest (CD4^+^ in this illustration). Subsequently, sequential gates were applied to identify markers of differentiation (CD45RA, CD197, CD27 and CD28), senescence (CD57) and exhaustion (CD57 and CD279(PD1)). Tn is identified as CD45RA^+^CD197^+^CD27^+^CD28^+^; Tcm are CD45RA^-^CD197^+^CD28^+^CD27^+/-^; Tem are CD45RA^-^CD197^-^CD28^-^CD27^+/-^; Te are CD45RA^+^CD197^-^CD28^-^CD27^-^CD57^+^. Tn, Naïve T-cells; Tcm, T-central memory; Tem, T-effector memory; Te, T-effector.

**Figure S5**

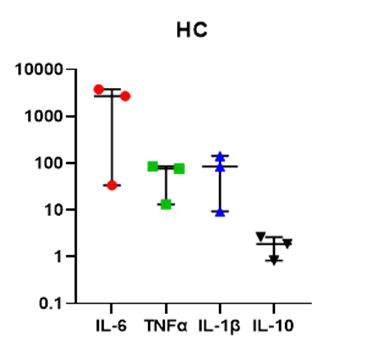

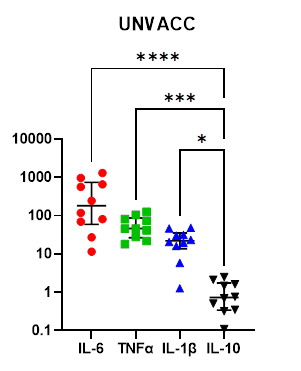

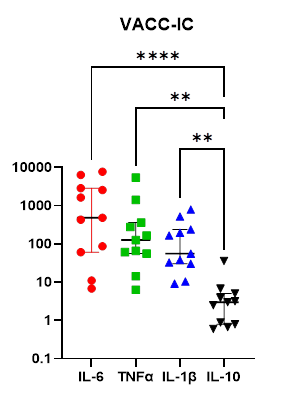

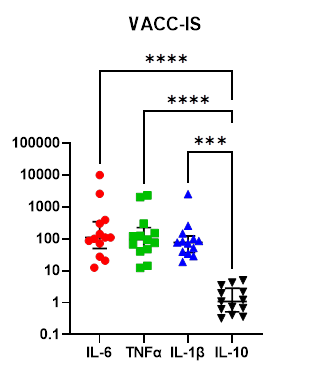

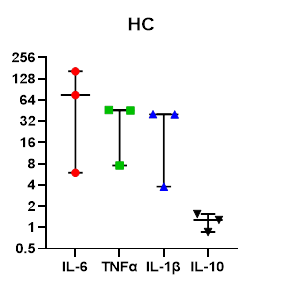

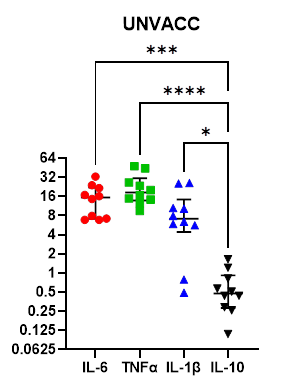

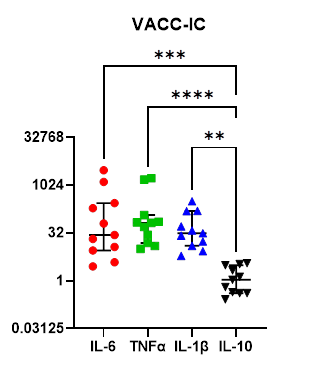

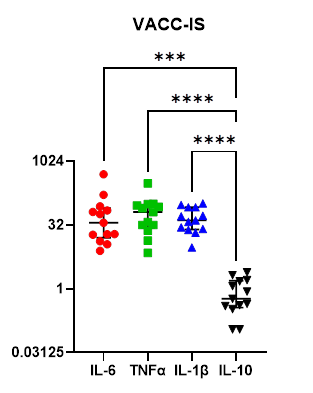

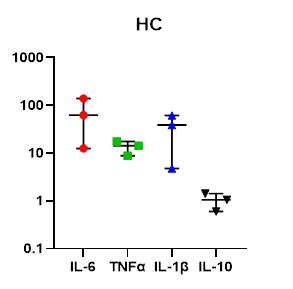

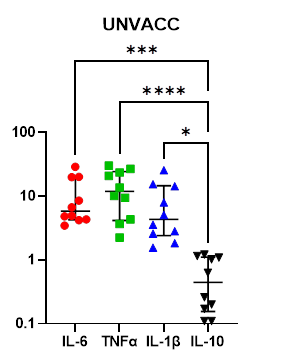

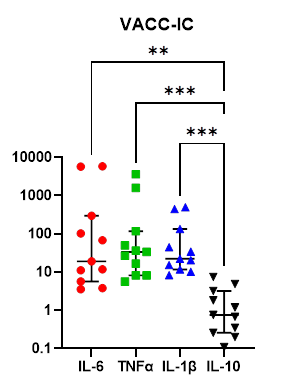

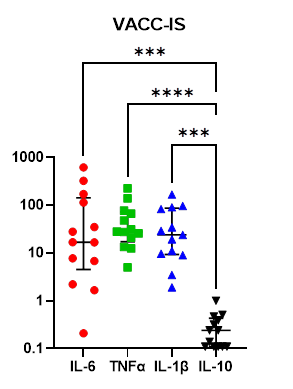

**A**

**B**

**C**

**pg/ml**

**Figure S5. Cytokine profile of each cohort post-SARS-CoV-2 peptide stimulation. Multiplex cytokine analysis was performed using supernatant of study participants PBMCs after S-peptide (A), NC-peptide (B) and MN- peptide (C) stimulation. Dot plots provide a comparative representative of** HC (n=3), UNVACC (n=10), VACC-IC (n=11) and VACC-IS (n=13) **cytokine profile, expressed as pg/ml, following SARS-CoV-2 peptide stimulation.** Two samples from UNVACC (n=1) and VACC-IC (n=1) were excluded due to laboratory technical error. Individual data points are shown here as scatter dot-plots with median (centre bar) and IQR (upper and lower bars). Statistical analyses were determined using nonparametric Kruskal-Wallis test with Dunn’s post-hoc test for multiple comparisons. **P*<0.05, ***P*<0.01, ****P*<0.001, *****P*<0.0001. HC, Healthy infection-naïve; UNVACC, unvaccinated convalescent; VACC-IC, vaccinated immunocompetent; VACC-IS, vaccinated immunosuppressed.
